# Assessment of psychometric validity and cross-cultural differential item functioning of the Doleur Neuropatique en 4 questions interview and the pain domain of the Western Ontario and McMaster Universities Osteoarthritis Index

**DOI:** 10.1101/2025.09.24.25336543

**Authors:** Jens Laigaard, Saber Muthanna Saber Aljuboori, Søren Overgaard, Karl Bang Christensen

## Abstract

**Background:** Randomised trials and meta-evidence increasingly rely on patient-reported outcome measures (PROMs). Many PROMs are applied in languages and settings that differ from the original target population. However, translation of PROMs poses a threat to their construct validity, including issues with cross-cultural adaptation

**Objective:** This study aims to assess the psychometric validity of the Danish versions of the WOMAC pain domain and the DN4i, including cross-cultural differential item functioning.

**Methods:** The study is based on a large dataset of 12-to 18-month pain outcomes after primary total hip arthroplasty (THA), total knee arthroplasty (TKA), or unicompartmental knee arthroplasty (UKA) for osteoarthritis (ClinicalTrials.gov identifiers NCT05845177 and NCT05900791). In addition to this dataset, we will apply for international data in similar populations to assess the cross-cultural differential item functioning.

The assessed instruments are the 5-item Western Ontario and McMaster Universities Osteoarthritis Index (WOMAC) pain domain (Likert-scale, version 3.1) and the 7-item Doleur Neuropatique en 4 questions interview (DN4i) (Danish translation from www.mapi-trust.org). We will evaluate if the data fit a congeneric measurement model, i.e. a model that assumes that the set of observed items all measure the same underlying latent factor. This evaluation of construct validity is done using Item Response Theory (IRT) and Confirmatory Factor Analysis (CFA).

**Perspective:** The results for WOMAC pain domain and DN4i will be reported in two separate reports, which are submitted for publication in peer-reviewed journals. We will seek to make the reports freely available, either by open-access publication or through publication on a preprint server, e.g. www.medrxiv.org.

## Introduction

Randomised trials and meta-evidence increasingly rely on patient-reported outcome measures (PROMs), which are often criticised for lack of validity. Most of the widely used PROMs are developed in English and then translated to other languages. This may affect the construct validity, i.e. that the PROM measures what it claims to measure.(1) Furthermore, issues with cross-cultural adaptation may also appear, i.e. when the translated PROMs fail to capture the cultural nuances and context-specific meanings, leading to potential biases and inaccuracies in the reported outcomes. For example, when using the Western Ontario and McMaster Universities Osteoarthritis Index (WOMAC) pain domain, the term ‘walking on a flat surface’ may be dissimilar between patients living in The Netherlands and patients living in the mountainous regions of Switzerland.(2) Such systematic bias can affect the accuracy, responsiveness, and confidence of trial results.(3) Local and cross-cultural validation of commonly used outcomes is therefore needed.

The WOMAC pain domain is a commonly applied outcome measure, particularly in chronic pain research.(2,4) The original English version has been validated and translated numerous times,(5) but the Danish version has not yet been validated. The French Doleur Neuropathique en 4 questions interview (DN4i) was developed to differentiate between patients with somatic and neurological tissue injuries.(6) It is also validated and translated extensively,(7,8) and can be used to identify patients who may have neuropathic pain.(9) However, many translations of the DN4i have not been validated, including the Danish (eprovide.mapi-trust.org/instruments/neuropathic-pain-4-questions).

In this study, we aim to assess the psychometric validity of the Danish versions of the WOMAC pain domain and the DN4i, including cross-cultural differential item functioning.

## Methods

### Data source

In 2023, the author group undertook two surveys of measures of pain and satisfaction 12-18 months after primary THA, TKA or UKA. The surveys were registered at ClinicalTrials.gov (identifiers NCT05845177 and NCT05900791) and listed at the Capitol Region of Denmark’s research (identifiers P-2022-933 and P-2023-4). The primary study populations were patients operated for primary osteoarthritis, namely 1148 UKA patients (870 respondents), 3086 TKA patients (2172 respondents), and 2777 THA patients (2031 respondents). However, a smaller number of patients operated for non-primary osteoarthritis were also surveyed. The median time from surgery to response was 17 months (interquartile range (IQR) 16-18 months) for THA patients, and 13 months (IQR 12-14 months) for UKA and TKA patients. Among other outcomes, the surveys included the following:

- **The 5-item Western Ontario and McMaster Universities Osteoarthritis Index (WOMAC) pain domain (Likert-scale, version 3.1)**.(2) *What amount of knee pain have you experienced the last week during the following activities?* a) *Walking on a flat surface, b) Going up or down stairs, c) At night while in bed, d) Sitting or lying, e) Standing upright*. Response options: None; Mild; Moderate; Severe; Extreme. Each of the five items is scored from 0 to 4. These scores are added to a 0-20 sum score, with a higher score indicating worse pain.
- **The 7-item Doleur Neuropatique en 4 questions interview (DN4i) (Danish translation from www.mapi-trust.org)**.(6,9) The DN4i assesses pain characteristics: *Does the pain have one or more of the following characteristics? a) Burning, b) Painful cold, c) electric shocks*, and associated symptoms: *Is the pain associated with one or more of the following symptoms in the same area? a) Tingling, b) Pins and needles, c) Numbness, d) Itching)*. Response options: Yes; No. Each of the seven items is scored from 0 to 1. The scores are added to a 0-7 sum score, with a higher score indicating more symptoms.

In addition to the data from the authors’ previous survey studies, we will also use data from published studies, that were conducted outside Denmark, where the included patients resembled ours, i.e. assessments of long-term outcomes after total hip arthroplasty (THA), total knee arthroplasty (TKA), and unicompartmental knee arthroplasty (UKA).

### Missing data

We will apply multiple imputation to impute missing item responses, however empty rows will be omitted from the dataset. We will use chained equations to generate the imputation-enriched dataset using predictive mean matching with the mice package in R (R Core Team, 2023).(10) The imputation model will include both demographic (baseline) and other outcome variables.

### Psychometric validation

The variables used for psychometric validation in this study are the instrument items (DN4i or WOMAC pain domain) and variables assessed for differential item functioning, e.g. data source (culture), sex and age. For this study, we will use only complete instrument responses, i.e. we will not employ any imputation. The psychometric validation evaluates reliability, i.e.,

1. **floor or ceiling effect**, i.e. if many respondents report the lowest or highest possible sum score, respectively. The flooring or ceiling effect is considered substantial if more than 15% of respondents have reported the lowest/highest possible score.
2. the distribution of sum scores, as evaluated with a histogram.
3. if the Cronbach’s alpha suggests high internal reliability (i.e. α ≥ 0.70). We chose to also assess Cronbach’s alpha because it is a widely used and interpretable index of reliability for reflective scales. However, with the more detailed evaluation of construct validity using confirmatory factor analysis (CFA), Cronbach’s alpha may be considered superfluous. Next, we evaluate if the data fit a congeneric measurement model, i.e. a model that assumes that the set of observed items all measure the same underlying latent factor. This evaluation of construct validity is done using **item response theory (IRT**) and **CFA** testing
4. if the instrument is **unidimensional**, i.e. that all items of the instrument measure the same latent variable. This is evaluated using correlations, visualised in a correlation matrix (heatmap), and CFA based on the
  - **Chi**^**2**^**-test**, which tests if the model-implied covariance matrix is significantly different from the observed covariance matrix. Should be insignificant (p>0.05), although biased against large datasets.
  - **Comparative Fit Index (CFI)** (i.e. how well does the factor model fit, compared to a baseline model assuming no relationships among the items). Should be CFI≥0.95.
  - **Root Mean Square Error of Approximation (RMSEA**) which essentially measures how different the proposed factor model is from ‘perfect fit’ to the data. Should be RMSEA≤0.06
  - **Standardized Root Mean Square Residual (SRMR**) which measures how well the model-implied correlations match the actual (observed) correlations among the items. Should be SRMR≤0.08.
5. if there is a **monotonous relationship between the individual items and the latent variable**, i.e. the sum score. Each item is plotted against the sum of the remaining items (**rest-score plots)**. As the score of an individual item increases, the sum score should also increase, with no reversals or inconsistencies. Furthermore, estimated factor loadings from CFA are inspected.
6. if there is no **differential item functioning (DIF)**, i.e. at the same sum score, each items function similarly across culture/sex/etc (cross-cultural validity). We will use **Ordinal Logistic Regression DIF**, to evaluate uniform and non-uniform DIF for each item. Uniform DIF means, that one group scores better/worse across all levels of the latent trait. Non-uniform DIF means that the direction or magnitude of the item bias changes at different levels of the latent trait. We will control for multiple comparisons using the Benjamini-Hochberg procedure (false discovery rate < 0.05).
7. if there is **local independence**, i.e. once we control for the sum score, responses to different items should be statistically independent of each other. This is tested similarly to DIF, using Yen’s Q3 residual correlations, plotted to a Residual Correlation Matrix. Q3 residuals > 0.20 above the average often indicate local dependence.

Other domains of the COSMIN checklist (e.g., content validity, criterion validity, and responsiveness) are not addressed here, as these have either been evaluated in prior studies or are not feasible/applicable in this context. The primary focus of this study is on cross-cultural validity, specifically the validation of the translation and the assessment of differential item functioning (DIF) compared to other cultures

## Data Availability

The data used in these studies are available from their primary sources upon reasonable request. Please see the original publications for details.

## Tables and figures

**Table 1:** A table of participants’ demographics stratified by culture (i.e. data source), including the scores, missingness, and sum score of the instrument.

**Table 2:**
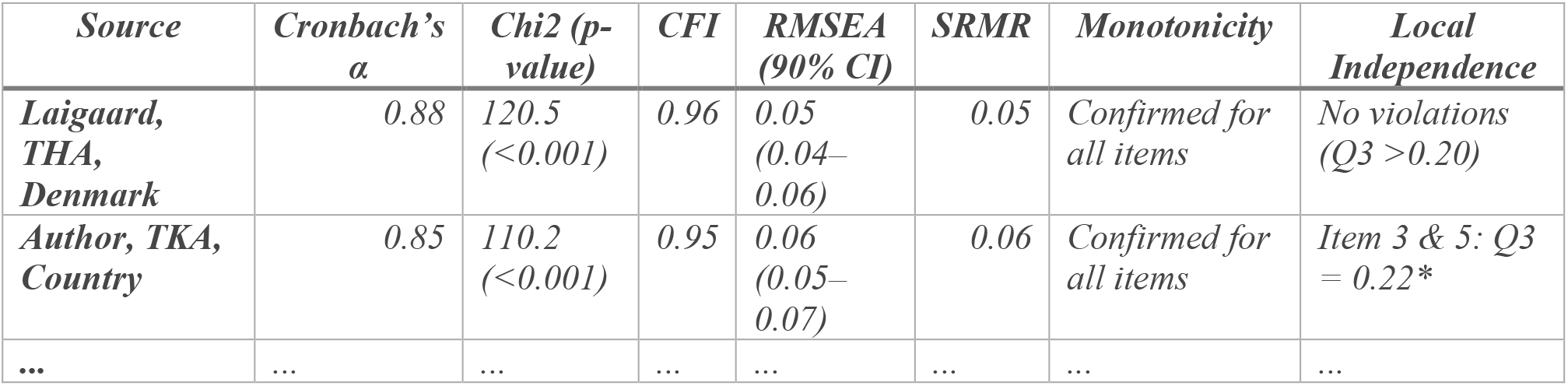
Construct validity indices, including Cronbach’s alpha and CFA fit indices (Chi^2^-test, CFI, RMSEA, SRMR, a note describing whether there is a monotonous relationship between the individual items and the latent variable, and a note on whether if there is local independence between items), stratified by culture (i.e. data source). For the notes, we will reference to the rest-score plots and residual correlation matrices where the information derives from (supplementary appendices). Mock table:

| *Source* | *Cronbach's*<br>*$\alpha$* | *Chi2 (p-*<br>*value)* | *CFI* | *RMSEA*<br>*(90% CI)* | *SRMR* | *Monotonicity* | *Local*<br>*Independence* |
| --- | --- | --- | --- | --- | --- | --- | --- |
| *Laigaard,*<br>*THA,*<br>*Denmark* | 0.88 | 120.5<br>($<0.001$) | 0.96 | 0.05<br>(0.04–<br>0.06) | 0.05 | Confirmed for<br>all items | No violations<br>($Q3 > 0.20$) |
| *Author, TKA,*<br>*Country* | 0.85 | 110.2<br>($<0.001$) | 0.95 | 0.06<br>(0.05–<br>0.07) | 0.06 | Confirmed for<br>all items | Item 3 & 5: $Q3 = 0.22^*$ |
| ... | ... | ... | ... | ... | ... | ... | ... |

**Figure 1.** Flow of participant response data

**Figure 2.** Histograms of instrument sum scores, stratified by source

**Figure 3.** A plot of DIF, for items that do express DIF. <u>For the DN4 interview</u>, the plot will be a probability plot of DIF with latent trait level (theta) as x-axis, and the probability of responding ‘yes’ to the item as y-axis, stratified by culture (i.e. data source). <u>For the WOMAC pain domain</u>, the plot will be a cumulative probability plot of differential item functioning with latent trait level (theta) as x-axis, and the cumulative probability of responding *None; Mild or less; Moderate or less; Severe or less; and Extreme or less*, as y-axis, stratified by culture (i.e. data source). Additional plots, such as correlation heatmaps, rest-score plots and residual correlation matrices will be attached as supplementary appendices.

## Knowledge dissemination

The results for WOMAC pain domain and DN4i will be reported in two separate reports, which are submitted for publication in peer-reviewed journals. We will seek to make the reports freely available, either by open-access publication or through publication on a preprint server, e.g. www.medrxiv.org.

## Ethical Considerations

The project is listed on the Capitol Region of Denmark’s research listing (p-2025-19417), which by delegation from The Danish Data Protection Agency approves the handling of confidential data for research. According to Danish legislation, approval from the national ethics committee is not required for studies that do not collect biological samples or impose interventions (Appendix 1)

Only Jens Laigaard will have access to the data, which are stored, handled and analysed pseudonymised at a logged and encrypted drive. For studies using data from the Danish National Prescription Registry, data are uploaded to Statistics Denmark’s secure online research platform in order to link the data.(11) This remote-access environment can only be accessed with the corresponding author’s unique digital signature.

## Appendix 1

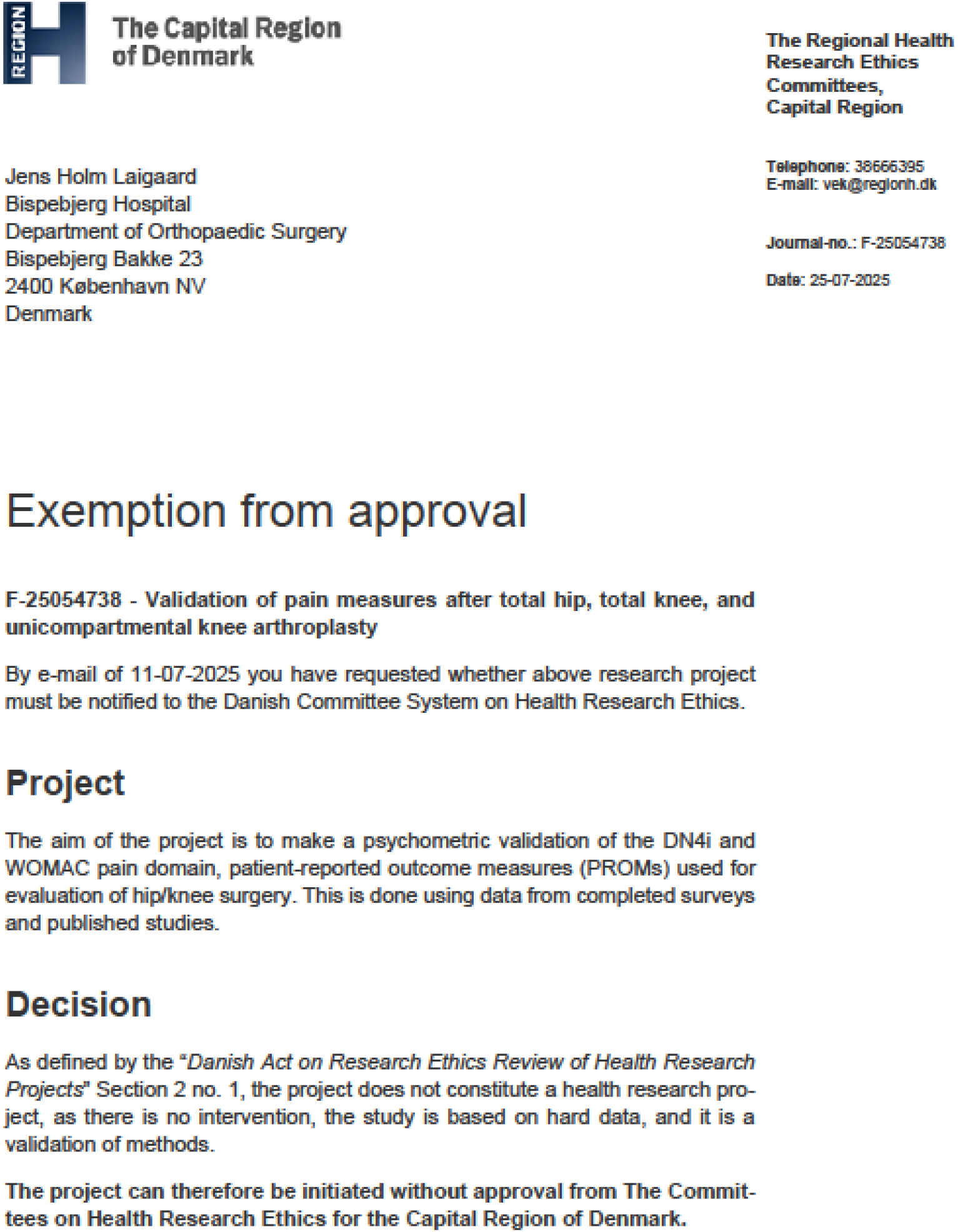

